# LOWERING CANCER-SPECIFIC MORTALITY RATES USING PROTEIN BIOMARKERS

**DOI:** 10.1101/2025.09.05.25335207

**Authors:** Borhan Mostafizi, Hamid RudsariKhosfekr, Ehsan Irajizad

**Affiliations:** James E Taylor; University of Texas MD Anderson Cancer Center

## Abstract

**Background:** Late-stage diagnosis remains a significant contributor to cancer mortality worldwide. Protein biomarkers have emerged as critical tools in both early detection and monitoring. This study investigates whether specific protein biomarkers can be leveraged not only to detect cancers at earlier stages but to reverse tumor progression toward more treatable stages, ultimately reducing mortality.

**Methods:** We evaluated 39 protein biomarkers across eight prevalent cancer types: breast, colorectal, esophageal, liver, lung, ovarian, pancreatic, and stomach. Biomarker performance was assessed using receiver operating characteristic (ROC) curve analysis to calculate the area under the curve (AUC) and sensitivities at 90% specificity across cancer stages I through III. The highest-performing biomarkers were subsequently used to estimate potential stage-shift effects and the corresponding reduction in mortality.

**Results:** Specific protein biomarkers demonstrated high predictive accuracy and strong potential for stage shifting across multiple cancers, which in turn may help reduce mortality. In breast cancer, a combination of prolactin and IL-8 achieved a 48.2% stage shift, translating into a 33.7% reduction in mortality. For colorectal cancer, integrating OPN and IL-8 led to a 68.7% stage shift, contributing to a 26.8% reduction in mortality. In esophageal cancer, OPN, HGF, and GDF15 shifted 65.4% of late-stage cases into earlier stages, resulting in a 12.4% mortality reduction. For liver cancer, Endoglin, HGF, and OPN together shifted 63% of late-stage cases to earlier stages, yielding a 4.4% reduction in mortality. In lung cancer, prolactin showed notable performance, shifting 65.1% of late-stage cancers to early stage, contributing to a 19% mortality reduction. Ovarian cancer demonstrated the greatest benefit, where prolactin and CA-125 shifted 79.1% of late-stage cancers to earlier stages, leading to a 50.3% decrease in projected deaths. In pancreatic cancer, TIMP-2 and CA19-9 shifted 81.1% of cases to earlier stages, helping lower mortality by 9.3%. Finally, in stomach cancer, OPN shifted 63.1% of late-stage cases to earlier stages, reducing mortality by 16.4%.

**Conclusions:** Protein biomarkers offer a promising path not only for early detection but also for mortality reduction. This study highlights the potential of biomarker-driven strategies to reduce cancer mortality through stage regression, particularly in common and lethal cancers.

**Impact:** These findings underscore the potential for integrating biomarker-guided interventions into cancer care, offering a new paradigm in personalized oncology focused on shifting the stage landscape toward improved survival.

## Introduction

Cancer remains the second leading cause of death worldwide, accounting for approximately 10 million deaths annually [1]. Every year, upwards of two million people develop cancer [10]. Despite advancements in screening, diagnostics, and strategies, the prognosis for many patients remains poor due to late-stage detection. Many cancer cases are detected at an advanced stage, when treatment options are limited [5].

Protein biomarkers, which are biological molecules present in blood, tissues, or other bodily fluids, have become increasingly important in the landscape of oncology [2]. Recent studies have demonstrated the promise of protein biomarkers in early cancer detection. Exact Sciences has developed multi-marker, blood-based assays for early colorectal and pan-cancer detection, including the Cologuard® test and its acquisition of Thrive Earlier Detection [3]. Their work is now in the late stages of commercialization, with tests already available in clinical settings for colorectal cancer and multi-cancer detection in development. There have also been developments in new technologies that have enabled the simultaneous detection of multiple cancers with high specificity, making “universal cancer screening” a possibility [12].

A study by Cohen et al. introduced CancerSEEK, a multi-analyte blood test that combines measurements of circulating proteins and tumor-derived DNA mutations to detect eight common cancers with high sensitivity and specificity [7]. The test demonstrated the ability to localize tumors and detect them at early stages, offering a minimally invasive method for early diagnosis [8].

Grail’s Galleri test uses cell-free DNA (cfDNA) and advanced methylation sequencing to detect more than 50 cancer types from a single blood draw. The technology identifies cancer-specific methylation patterns, providing both detection and tissue-of-origin information. Grail has already launched Galleri in the commercial market, and it is being evaluated in large-scale prospective trials such as the PATHFINDER and NHS-Galleri studies, making it a very commercially advanced multi-cancer early detection (MCED) test [8]. Including all the other studies, there’s solid reason that supports the potential for novel, multi-cancer early detection (MCED) tests that can be applied to population-wide cancer screening [13].

A promising approach in cancer management is the concept of stage shifting: altering the biological trajectory of tumors to earlier stages where prognosis is significantly better [19,31]. Leveraging high-performing protein biomarkers to induce or model such shifts could offer a transformative avenue for reducing cancer mortality [20,21,22,24,30,33]. Many protein biomarkers have been recently found to be specifically concentrated around the area of the cancer. By targeting those proteins, the idea of earlier detection of cancers seems promising [32]. Protein biomarkers are molecules that indicate the start or happening of specific processes or conditions inside an organism [23].

In this study, we investigate the utility of 39 protein biomarkers across eight common cancer types. Using ROC curve analyses and publicly available datasets, we identify top-performing biomarkers based on their predictive power and model their impact on stage distribution and cancer-specific mortality.

## Materials and Methods

The dataset used in this study is from the publicly available CancerSEEK dataset, which includes blood test results from 1,818 individuals. These results measure 39 different protein biomarkers for people with one of eight types of cancer: breast, colorectal, esophageal, liver, lung, ovarian, pancreatic, and stomach cancer [7]. The dataset includes information about which stage of cancer each person had (Stages I, II, or III) and their characteristics. These details allowed us to study how well each protein could detect different cancers at early stages. The specific details of the CancerSeek dataset are in Table 1.

**Table 1.**
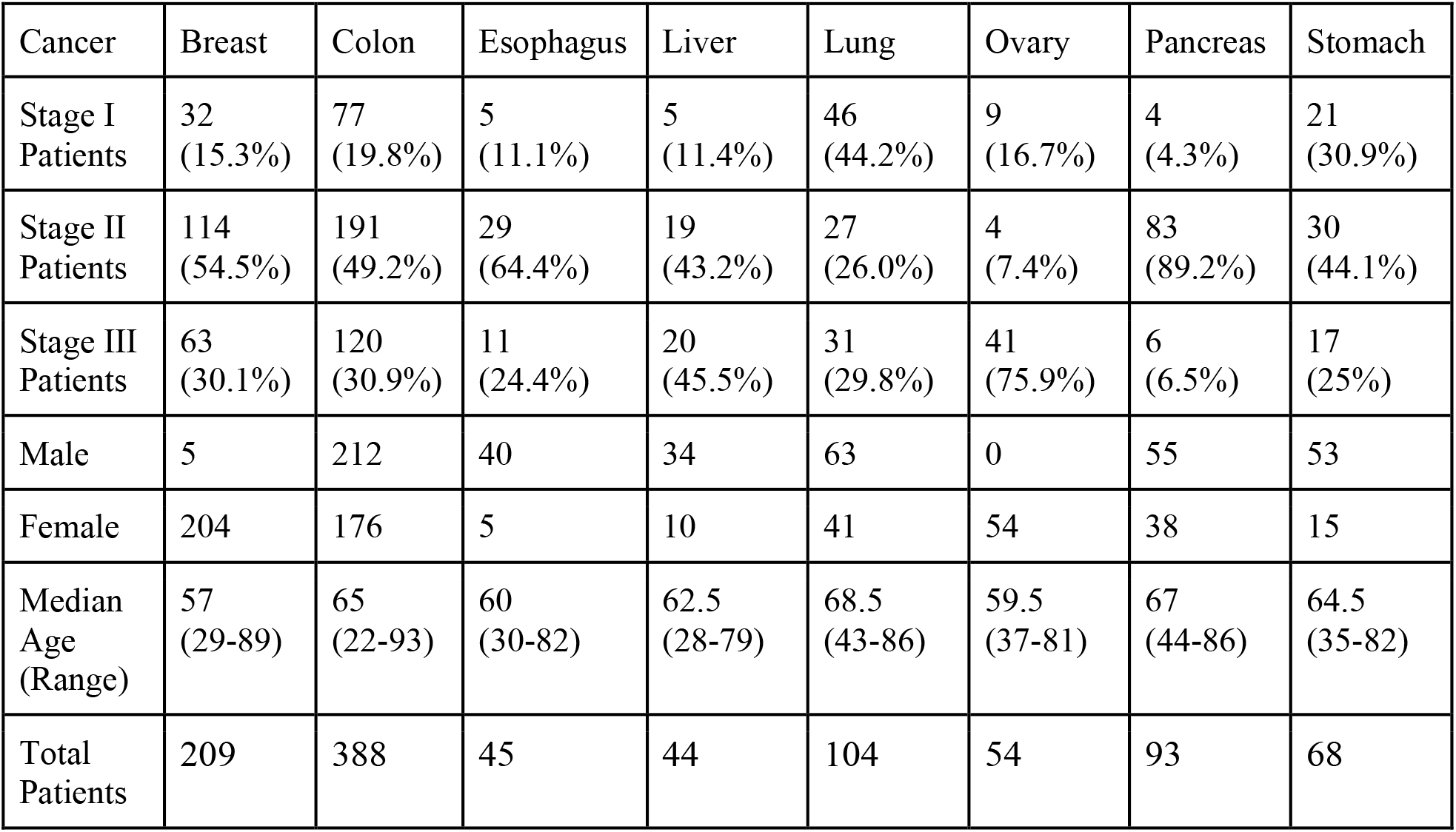
Characteristics and numbers of the patients.

| Cancer | Breast | Colon | Esophagus | Liver | Lung | Ovary | Pancreas | Stomach |
| --- | --- | --- | --- | --- | --- | --- | --- | --- |
| Stage I Patients | 32<br>(15.3%) | 77<br>(19.8%) | 5<br>(11.1%) | 5<br>(11.4%) | 46<br>(44.2%) | 9<br>(16.7%) | 4<br>(4.3%) | 21<br>(30.9%) |
| Stage II Patients | 114<br>(54.5%) | 191<br>(49.2%) | 29<br>(64.4%) | 19<br>(43.2%) | 27<br>(26.0%) | 4<br>(7.4%) | 83<br>(89.2%) | 30<br>(44.1%) |
| Stage III Patients | 63<br>(30.1%) | 120<br>(30.9%) | 11<br>(24.4%) | 20<br>(45.5%) | 31<br>(29.8%) | 41<br>(75.9%) | 6<br>(6.5%) | 17<br>(25%) |
| Male | 5 | 212 | 40 | 34 | 63 | 0 | 55 | 53 |
| Female | 204 | 176 | 5 | 10 | 41 | 54 | 38 | 15 |
| Median Age (Range) | 57<br>(29-89) | 65<br>(22-93) | 60<br>(30-82) | 62.5<br>(28-79) | 68.5<br>(43-86) | 59.5<br>(37-81) | 67<br>(44-86) | 64.5<br>(35-82) |
| Total Patients | 209 | 388 | 45 | 44 | 104 | 54 | 93 | 68 |

For this research, we assumed that the Stage III patients include Stage IV patients. To evaluate the role of protein biomarkers in cancer progression, we first calculated the presence of each protein in the tumor stages of interest (Stages I-III) across the eight types of cancer. Biosensors and detectors that would be necessary for this hypothesized method to occur have been recently made and can successfully detect cancer protein biomarkers with ultrahigh sensitivity [27, 28, 29]. The biomarkers under investigation were: AFP, Angiopoietin-2, AXL, CA-125, CA15-3, CA19-9, CD44, CEA, CYFRA21-1, DKK1, Endoglin, FGF2, Follistatin, Galectin-3, G-CSF, GDF15, HE4, HGF, IL-6, IL-8, Kallikrein, Leptin, Mesothelin, Midkine, Myeloperoxidase, NSE, OPG, OPN, PAR, Prolactin, sEGFR, sFas, SHBG, sHER2, sPECAM-1, TGFa, Thrombospondin-2, TIMP-1, and TIMP-2.

To understand the predictive power of each biomarker, we utilized data from the CancerSEEK project. We calculated each biomarker’s Area Under the Curve (AUC) by creating a Receiver Operating Characteristic (ROC) curve for each tumor stage. The ROC curve is used both to represent the overall performance of a diagnostic test at cut-off points and to determine the optimal cut-off value for diagnosing a disease [25]. The AUC is a statistical measure used to quantify the overall performance of a diagnostic test and can be interpreted as the average value of sensitivities for all possible specificities [25]. The biomarkers with the greatest AUC values are aggregated into Table 2. All AUC values for each protein biomarker in a specific cancer type and stage are in GitHub, which is linked in the ‘Supplements’ part of the paper. We then found the sensitivity of each of the highest-performing proteins at a 90% specificity threshold. Sensitivity is the probability of a positive test given that the person has cancer, while specificity is the probability of a negative test given that the person does not have cancer [9]. The sensitivities of the best proteins are aggregated into Table 3.

**Table 2.** Summary of the protein biomarkers with the greatest AUC value for each cancer and its corresponding stage.

| CANCER TYPE AND STAGE | Highest performing Biomarker (AUC) | CANCER TYPE AND STAGE | Highest performing Biomarker (AUC) |
| --- | --- | --- | --- |
| Breast Cancer I | Prolactin (0.7423414) | Lung Cancer I | Prolactin (0.8889216) |
| Breast Cancer II | IL-8 (0.7427567) | Lung Cancer II | Prolactin (0.9383780) |
| Breast Cancer III | IL-8 (0.8099050) | Lung Cancer III | Prolactin (0.8817337) |
| Colorectal Cancer I | OPN (0.8578626) | Ovarian Cancer I | Prolactin (0.9158456) |
| Colorectal Cancer II | IL-8 (0.8898557) | Ovarian Cancer II | Prolactin (1.0000000) |
| Colorectal Cancer III | IL-8 (0.8452586) | Ovarian Cancer III | CA-125 (0.9802655) |
| Esophageal Cancer I | OPN (0.8288177) | Pancreatic Cancer I | TIMP-2 (0.9592057) |
| Esophageal Cancer II | HGF (0.9492526) | Pancreatic Cancer II | CA19-9 (0.9252107) |
| Esophageal Cancer III | GDF15 (0.8562472) | Pancreatic Cancer III | CA19-9 (1.0000000) |
| Liver Cancer I | Endoglin (0.9330049) | Stomach Cancer I | OPN (0.9482172) |
| Liver Cancer II | HGF (0.9784159) | Stomach Cancer II | OPN (0.9157635) |
| Liver Cancer III | OPN (0.9886084) | Stomach Cancer III | OPN (0.9668212) |

**Table 3.** Sensitivities of the highest-performing biomarkers for each cancer and stage at 90% specificity.

| Cancer Type | Sensitivity in Stage I | Sensitivity in Stage II | Sensitivity in Stage III |
| --- | --- | --- | --- |
| Breast Cancer | 0.594 | 0.272 | 0.349 |
| Colon Cancer | 0.558 | 0.675 | 0.467 |
| Esophageal Cancer | 0.6 | 0.897 | 0.455 |
| Liver Cancer | 0.8 | 0.895 | 1 |
| Lung Cancer | 0.761 | 0.815 | 0.742 |
| Ovarian Cancer | 0.889 | 1 | 0.561 |
| Pancreatic Cancer | 1 | 0.819 | 1 |
| Stomach Cancer | 0.81 | 0.667 | 0.882 |

### Stage-shifting

To validate our findings, we applied our sensitivity values of proteins into Hubbell et al.’s equations [4]. We calculate the estimated stage shift as it has been proven to significantly lower mortality rates [11]. 4 necessary inputs are needed to successfully convert the protein’s sensitivity into the number of cases that could be detected earlier: protein sensitivity, stage-specific dwell time, screening interval, and a dataset. Using the values from the previous step, the dwell times stated by Hubbell in the “Fast” scenario described in Supplementary Figure S4, a typical 1-year screening interval, and the CancerSEEK database, we were able to use the equations. By adopting this model, we were able to estimate stage shift proportions under an intervention strategy guided by biomarker performance. Below is an overview of the simulation:

Marginal Sensitivity Calculation: For each stage *i*, the marginal sensitivity (Δs_i_) was calculated as the difference between the cumulative sensitivity at stage *i* and the cumulative sensitivity at the preceding stage (e.g., Δs_2_ = s_2_ − s_1_).

Slip Rate Estimation: Using the dwell time *D*_*i*_ for each stage, the probability of slipping past stage *i* without detection was calculated as *r*_*i*_ *= e^(−1/D*_*i*_).

Detection and Interception Modeling (specified for stage III patients):

1. Stage I detection: Patients detectable at Stage I were estimated as *N × Δs*_*1*_ *× (1 − r*_*1*_).
2. Slippage to Stage II: Patients who slipped from Stage I to Stage II were calculated as *N × Δs*_*1*_ *× r*_*1*_, added to those originally detectable at Stage II *(N × Δs*_*2*_).
3. Stage II detection: Intercepted patients at Stage II were estimated as *(total at II) × (1 − r*_*2*_), with the remainder slipping to Stage III.
4. Stage III detection: Patients remaining in Stage III were those who were not intercepted at earlier stages.

*N*: Number of Patients | *Δs*: Marginal Sensitivity | *r*: slip rate

Stage Shift Proportions: The number of patients intercepted at each earlier stage was divided by the original late-stage case count to determine the percentage of stage shifts (e.g., III → II, III → I). The resulting stage shift percentages are summarized in Table 4 and represent the idealized potential for early detection under biomarker-guided screening.

**Table 4.** Stage shift percentages from stage 3 to an earlier stage & stage 2 to stage 1 for each of the 8 cancer types.

| Cancer Type | III → II/I |
| --- | --- |
| Breast Cancer | 48.2% |
| Colon Cancer | 68.7% |
| Esophageal Cancer | 65.4% |
| Liver Cancer | 63% |
| Lung Cancer | 65.1% |
| Ovarian Cancer | 79.1% |
| Pancreatic Cancer | 81.1% |
| Stomach Cancer | 63.1% |

### Mortality reduction

Following the study by Owens et. al, we modeled the impact of hypothetical stage shifts on cancer mortality [6]. The underlying premise is based on the well-established observation that cancers detected at earlier stages have significantly lower mortality rates than those diagnosed at later stages. To validate the effectiveness of these hypothetical stage shifts, we applied a model that calculated the expected mortality rates after the stage shifts, using the formula:

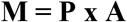

Where:

- M represents the expected mortality rate that has been reduced.
- A is the proportion of patients who shifted stages, as determined in the previous step.
- P is the surrogate endpoint (Pearson’s universal mortality rate; specific per cancer)

These surrogate mortality rates represent the average probability of death for a patient diagnosed at a late stage and were drawn from Owens’ published paper [6]. For this study, the following values were used: Breast Cancer-0.70; Colorectal Cancer-0.39; Esophageal Cancer-0.19; Liver Cancer-0.07; Lung Cancer-0.29; Ovarian Cancer-0.64; Pancreatic Cancer-0.12; Stomach Cancer-0.26.

By multiplying the proportion of patients expected to shift stages (A) with the baseline late-stage mortality (P) for each cancer, we estimated the number of deaths potentially averted through biomarker guided early detection. This approach assumes that the primary benefit of stage shifting lies in accessing earlier, more effective treatment options, which correspond to lower mortality risk. The resulting mortality reductions are summarized in Table 5.

**Table 5.** Estimated Mortality Reductions in Breast, Colon, Lung, and Ovarian Cancer.

| Cancer | Estimated Mortality Reduction |
| --- | --- |
| Breast Cancer | 33.7% |
| Colon/Rectum Cancer | 26.8% |
| Esophageal Cancer | 12.4% |
| Liver Cancer | 4.4% |
| Lung Cancer | 19.0% |
| Ovarian Cancer | 50.3% |
| Pancreatic Cancer | 9.3% |
| Stomach Cancer | 16.4% |

## Results

Analysis of the CancerSEEK dataset revealed that several protein biomarkers demonstrated strong discriminative performance across cancer types and stages. These high AUC values indicate that these biomarkers possess robust potential for detecting cancer at specific stages.

Table 2 summarizes the top-performing biomarkers by cancer type and stage. For instance, IL-8 was consistently elevated in breast and colorectal cancer. In lung cancer, Prolactin showed outstanding performance, achieving an AUC of 0.94 at stage II. Ovarian cancer exhibited near-perfect detection accuracy using Prolactin, while CA15-3 also performed well at stage III. Other high-performing biomarkers included OPN in breast, stomach, and colorectal cancers, and CA19-9 in pancreatic cancer. These consistent patterns suggest that a select group of biomarkers may serve as promising candidates for multi-cancer early detection strategies.

We then calculated the sensitivities of the proteins with the highest AUC value at a 90% specificity. The sensitivities are listed in Table 3. For the study, if a stage’s sensitivity was lower than that of the previous stage, the previous stage’s sensitivity value was used instead.

We then plugged the sensitivity values of the best biomarkers into the adjusted Hubbell simulation. The derived simulation converts protein sensitivity into stage shift percentages through state transition modeling (Table 3).

Using the stage-shift results and applying the simplified mortality reduction model (M = P × A) from Hubbell, we estimated reductions in cancer-specific mortality for the cancers. These estimates are shown in Table 5.

## Discussion

This study demonstrates the transformative potential of protein biomarkers in reducing cancer related mortality through early detection and stage shifting. By analyzing data from the CancerSEEK database and validating our findings with external modeling datasets, we identified high-performing biomarkers, such as Prolactin, OPN, IL-8, and CA19-9, that are not only consistently present across multiple cancer types but also possess strong predictive power, as reflected in their AUC values. These biomarkers serve as crucial indicators of cancer progression and offer promising targets for intervention aimed at shifting tumors to earlier, more treatable stages.

Breast Cancer demonstrated a 33.7% reduction in mortality rates following stage shifts. The high proportion of patients who could shift from Stage IV and III to II and I suggests that targeting biomarkers such as OPN and IL-8 could yield substantial clinical benefits. With earlier intervention, survival rates for breast cancer could be significantly improved, leading to a healthier population and a reduction in the burden on healthcare systems. This is especially important considering the prevalence of breast cancer globally and the current treatment challenges associated with advanced stages. The specific stage shift for breast cancer is modeled in Figure 2.a.

**Figure 1.**
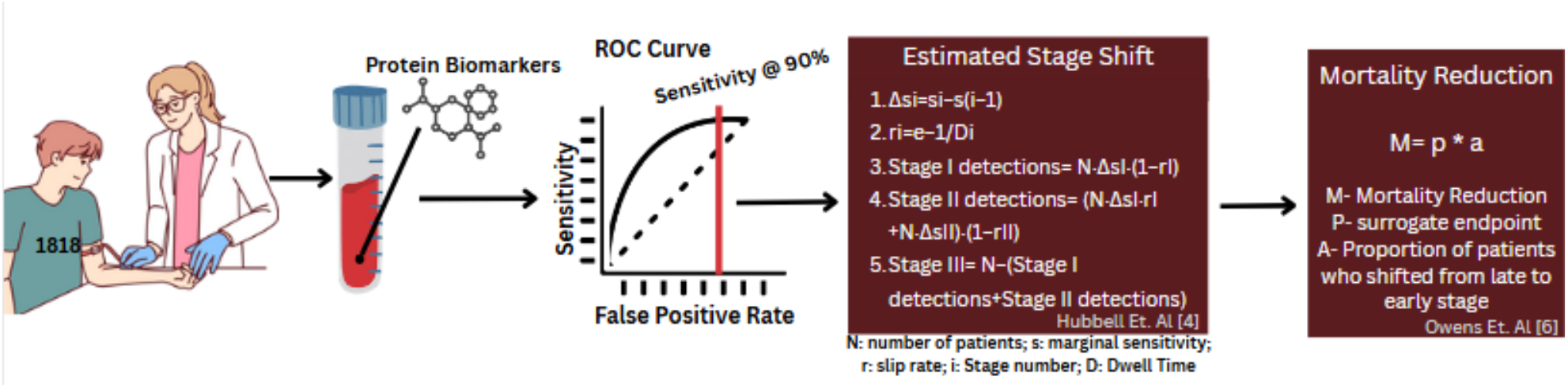
Overview of the research [35].

**Figure 2.**
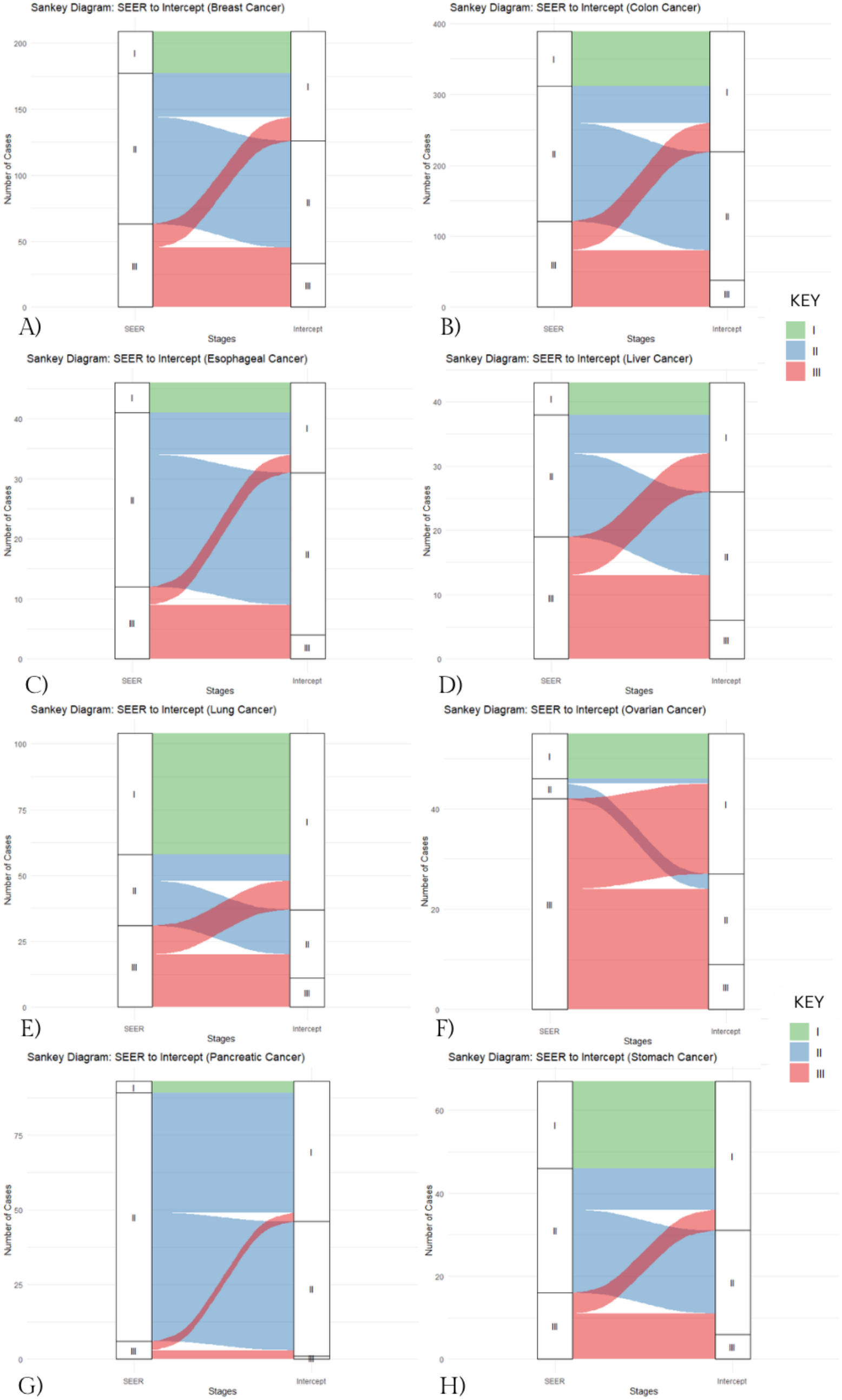
Model of the stage shift for each of the cancers.

In Colon/Rectum Cancer, we observed a 26.8% reduction in mortality rates. Although not as dramatic as the reduction seen in other cancers like ovarian, this still signifies a meaningful improvement in patient outcomes. By shifting patients from Stage III and IV to earlier stages, the cancer becomes more manageable, and patients experience better prognoses. The potential to delay cancer progression or reduce its severity is particularly crucial in colon cancer, where early detection and intervention are key to long term survival. Figure 2.b illustrates the specific stage shift for colon cancer.

Esophageal cancer demonstrated a 65.4% stage shift from a late stage towards an earlier stage, resulting in an estimated 12.4% reduction in mortality. Although this improvement is modest compared to other cancer types, it represents a meaningful step forward given the typically poor prognosis associated with esophageal cancer. Biomarker-driven strategies that enable earlier intervention may help convert esophageal cancer into a more treatable condition, ultimately enhancing survival rates and patient quality of life. The esophageal cancer stage shift is depicted in Figure 2.c.

In liver cancer, a 63% stage shift translates into a 4.4% reduction in cancer-specific mortality. Despite the relatively high stage-shift percentage, the mortality reduction remains low due to the generally poor survival rates even in earlier stages of liver cancer. Future efforts may need to combine early-stage identification with more effective technology to maximize survival benefits. Nevertheless, any shift away from late-stage presentation can still improve clinical management and reduce treatment burden. Figure 2.d presents the modeled stage shift for liver cancer.

Lung Cancer, with a reduction of 19%, demonstrates one of the most significant improvements in mortality rates after stage shifting. Lung cancer remains one of the leading causes of cancer-related deaths worldwide, and its typically late-stage diagnosis often results in poor survival rates. Targeting biomarkers like Prolactin and IL-8 could drastically improve survival by delaying progression to late stages. These findings underscore the importance of early-stage interventions in lung cancer, offering a potential breakthrough in how we treat this often aggressive disease. The modeled lung cancer stage shift is shown in Figure 2.e.

In the case of Ovarian Cancer, a remarkable 50.3% reduction in mortality rates is predicted. This drastic improvement highlights the power of protein biomarkers in shifting cancer stages, particularly for cancers like ovarian, which are often diagnosed at later stages due to their subtle symptomatology. By identifying and targeting biomarkers such as Prolactin, clinicians may be able to reverse disease progression and offer patients a better chance of survival. The substantial mortality rate reduction for ovarian cancer emphasizes the promise of precision medicine based on individual biomarker profiles. Figure 2.f displays the stage shift pattern specific to ovarian cancer.

Pancreatic cancer exhibited a high 81.1% stage shift, yet this only yielded a 9.3% mortality reduction, highlighting the stark reality of a disease known for its lethality across all stages. The modest reduction, despite dramatic stage migration, reflects the limited survival gains achievable with current treatments, even when pancreatic cancer is detected early [34]. Identifying highly predictive biomarkers for earlier detection, combined with novel treatment modalities, may be key to improving outcomes in this devastating cancer. The stage shift for pancreatic cancer is visualized in Figure 2.g.

Stomach cancer demonstrated a 63.1% shift from Stage III to earlier stages, leading to an estimated 16.4% mortality reduction. Although not as dramatic as seen in ovarian or lung cancer, this improvement is still clinically significant, especially given its nonspecific and late-presenting symptoms. Earlier diagnosis could enable more effective surgical interventions and tailored treatment regimens. The results suggest that enhancing biomarker-based detection tools for stomach cancer may help convert it into a more survivable condition, particularly in high-risk populations. Figure 2.h provides a model of the ovarian cancer stage shift.

## Conclusion

Our findings highlight that specific biomarkers, when targeted, could effectively shift cancer stages backward, leading to substantial reductions in mortality rates. Furthermore, this approach not only demonstrates the potential for improving survival outcomes but also emphasizes the broader application of protein biomarkers in cancer treatment strategies.

Beyond mortality reduction, the application of protein biomarkers in clinical settings holds promise for advancing personalized medicine. By tailoring interventions based on an individual’s biomarker profile, clinicians may be able to intervene earlier, reduce treatment burden, and improve patient quality of life. This research establishes a foundational model for integrating biomarker-guided strategies into mainstream oncology, highlighting the need for further clinical trials and the development of targeted treatment protocols that capitalize on the biological specificity of these proteins.

While our results are promising, several key limitations and unanswered questions remain. First, the biological mechanisms by which biomarkers like Prolactin or IL-8 might induce backward stage shifts require further study [15]. Second, we did not address the potential risks of targeting these biomarkers, including off-target effects or interference with normal physiological functions, particularly since many biomarkers serve roles in healthy tissues [16]. Thirdly, while multi-cancer early detection (MCED) tests offer the promise of shifting diagnoses to earlier, more treatable stages, they also introduce the risk of overdiagnosis and false positives, particularly for cancers that may never progress clinically [14]. In addition, a very scarce amount of methods using protein biomarkers have been evaluated with real patient samples, which is key to establishing clinical sensitivity and selectivity [26].

Furthermore, although the stage shift modeling was based on neutralistic assumptions, its real world applicability may be limited [4,17]. A sensitivity analysis using more conservative clinical parameters could help assess the robustness of these predictions. In addition, our validation relied on simulated data rather than an independent real-world patient cohort, which limits the external generalizability of the findings [18]. Finally, while we demonstrate strong predictive performance of certain biomarkers, the translational feasibility of developing diagnostics based on these proteins was not explored. Future research should focus on bridging this translational gap and exploring whether similar results can be observed in other cancer types beyond the eight analyzed here.

In conclusion, our research provides compelling evidence that protein biomarkers can revolutionize cancer care by enabling earlier detection and enabling stage regression. The significant mortality reductions observed across multiple cancer types affirm the clinical relevance of our findings and underscore the urgent need to translate biomarker discoveries into actionable treatment strategies. As the field moves toward more precise and personalized approaches, protein biomarkers will be central to reshaping cancer prognosis, reducing mortality, and ultimately saving lives.

## Data Availability

Data is publicly available elsewhere

https://pubmed.ncbi.nlm.nih.gov/29348365/

## Supplementals

https://github.com/BorhanMos/ProteinBiomarkerStageShiftAndMortalityModel

## Citations

1. American Cancer Society. “The Global Cancer Burden | American Cancer Society.” http://www.cancer.org, 2024, http://www.cancer.org/about-us/our-global-health-work/global-cancer-burden.html.

2. Melone, Mary. “The Role of Oncology Biomarkers in Personalizing Hematology Treatment Plans.” Worldwide Clinical Trials, 14 Aug. 2024, http://www.worldwide.com/blog/2024/08/oncology-biomarkers-in-hematology-treatment/.

3. “Exact Sciences to Acquire Thrive Earlier Detection, Becoming a Leader in Blood-Based, Multi-Cancer Screening.” Exactsciences.com, 2018, investor.exactsciences.com/investor-relations/press-releases/press-release details/2020/Exact-Sciences-To-Acquire-Thrive-Earlier-Detection-Becoming-A-Leader-In-Blood-Based-Multi-Cancer Screening/default.aspx.

4. Hubbell, Earl, et al. “Modeled Reductions in Late-Stage Cancer with a Multi-Cancer Early Detection Test.” Cancer Epidemiology and Prevention Biomarkers, vol. 30, no. 3, 1 Mar. 2021, pp. 460–468, cebp.aacrjournals.org/content/30/3/460, 10.1158/1055-9965.EPI-20-1134.

5. Crosby, David, et al. “Early Detection of Cancer.” Science, vol. 375, no. 6586, 18 Mar. 2022, http://www.science.org/doi/10.1126/science.aay9040, 10.1126/science.aay9040.

6. Owens, Lukas, et al. “Stage Shift as an Endpoint in Cancer Screening Trials: Implications for Evaluating Multicancer Early Detection Tests.” Cancer Epidemiology, Biomarkers & Prevention, vol. 31, no. 7, 27 Apr. 2022, pp. 1298–1304, 10.1158/1055-9965.epi-22-0024.

7. Cohen, Joshua D., et al. “Detection and Localization of Surgically Resectable Cancers with a Multi-Analyte Blood Test.” Science, vol. 359, no. 6378, 18 Jan. 2018, pp. 926–930, http://www.ncbi.nlm.nih.gov/pmc/articles/PMC6080308/, 10.1126/science.aar3247.

8. Liu, M.C., et al. “Sensitive and Specific Multi-Cancer Detection and Localization Using Methylation Signatures in Cell-Free DNA.” Annals of Oncology, vol. 31, no. 6, Mar. 2020, 10.1016/j.annonc.2020.02.011.9.

9. “Cancer Screening Overview.” National Cancer Institute, Cancer.gov, 2011, http://www.cancer.gov/aboutcancer/screening/hp-screening-overview-pdq.

10. American Cancer Society. “Cancer Facts & Figures 2025.” Cancer.org, 2025, http://www.cancer.org/research/cancer-factsstatistics/all-cancer-facts-figures/2025-cancer-facts-figures.html.

11. Wever, Elisabeth M., et al. “How Does Early Detection by Screening Affect Disease Progression?” Medical Decision Making, vol. 31, no. 4, 15 Mar. 2011, pp. 550–558, 10.1177/0272989×10396717.

12. Ahlquist, David A. “Universal Cancer Screening: Revolutionary, Rational, and Realizable.” Npj Precision Oncology, vol. 2, no. 1, 29 Oct. 2018, pp. 1–5, http://www.nature.com/articles/s41698-018-0066-x, 10.1038/s41698-018-0066-x.

13. Srivastava, Sudhir, and Sam Hanash. “Pan-Cancer Early Detection: Hype or Hope?” Cancer Cell, vol. 38, no. 1, July 2020, pp. 23–24, 10.1016/j.ccell.2020.05.021. Accessed 8 Apr. 2021.

14. Klein, E. A., et al. “Clinical Validation of a Targeted Methylation-Based Multi-Cancer Early Detection Test Using an Independent Validation Set.” Annals of Oncology, vol. 32, no. 9, 1 Sept. 2021, pp. 1167–1177, http://www.annalsofoncology.org/article/S0923-7534(21)02046-9/fulltext, 10.1016/j.annonc.2021.05.806.

15. Paliouras, Miltiadis, and Eleftherios P. Diamandis. “Androgens Act Synergistically to Enhance Estrogen-Induced Upregulation of Human Tissue Kallikreins 10, 11, and 14 in Breast Cancer Cells via a Membrane Bound Androgen Receptor.” Molecular Oncology, vol. 1, no. 4, 9 Jan. 2008, pp. 413–424, 10.1016/j.molonc.2008.01.001. Accessed 15 Mar. 2022.

16. Poste, George. “Bring on the Biomarkers.” Nature, vol. 469, no. 7329, Jan. 2011, pp. 156–157, 10.1038/469156a.

17. Etzioni, Ruth, et al. “The Case for Early Detection.” Nature Reviews Cancer, vol. 3, no. 4, Apr. 2003, pp. 243–252, 10.1038/nrc1041.

18. He, Zhe, et al. “Clinical Trial Generalizability Assessment in the Big Data Era: A Review.” Clinical and Translational Science, vol. 13, no. 4, 1 July 2020, pp. 675–684, pmc.ncbi.nlm.nih.gov/articles/PMC7359942/, 10.1111/cts.12764.

19. Yang, Ching-Yao. Stage Shift Improves Lung Cancer Survival: Real-World Evidence. journal of thoracic oncology, 10 Apr. 2020, http://www.jto.org/article/S1556-0864(22)01591-X/fulltext#:~:text=PS01.02%20National%20Lung%20Cancer,in%20turn%2C%20may%20improve%20survival.

20. Alexandre Mezentsev, et al. “A Comprehensive Review of Protein Biomarkers for Invasive Lung Cancer.” Current Oncology, vol. 31, no. 9, 23 Aug. 2024, pp. 4818–4854, http://www.mdpi.com/1718-7729/31/9/360, 10.3390/curroncol31090360.

21. Geary, Bethany, et al. “Discovery and Evaluation of Protein Biomarkers as a Signature of Wellness in Late-Stage Cancer Patients in Early Phase Clinical Trials.” Cancers, vol. 13, no. 10, 18 May 2021, pp. 2443–2443, pmc.ncbi.nlm.nih.gov/articles/PMC8157875/, 10.3390/cancers13102443.

22. Barker, Anna D, et al. “An Inflection Point in Cancer Protein Biomarkers: What Was and What’s Next.” Molecular & Cellular Proteomics, vol. 22, no. 7, 1 July 2023, pp. 100569–100569, 10.1016/j.mcpro.2023.100569.

23. Biotechnology, Nautilus. “What Are Protein Biomarkers?” Nautilus Biotechnology, 18 Apr. 2023, http://www.nautilus.bio/blog/what-are-protein-biomarkers/.

24. Boschetti, Egisto, et al. “Protein Biomarkers for Early Detection of Diseases: The Decisive Contribution of Combinatorial Peptide Ligand Libraries.” Journal of Proteomics, vol. 188, 30 Sept. 2018, pp. 1–14, http://www.sciencedirect.com/science/article/abs/pii/S1874391917302853, 10.1016/j.jprot.2017.08.009.

25. Nahm, Francis Sahngun. “Receiver Operating Characteristic Curve: Overview and Practical Use for Clinicians.” Korean Journal of Anesthesiology, vol. 75, no. 1, 1 Feb. 2022, pp. 25–36, http://www.ncbi.nlm.nih.gov/pmc/articles/PMC8831439/, 10.4097/kja.21209.

26. Rusling, James F., et al. “Measurement of Biomarker Proteins for Point-of-Care Early Detection and Monitoring of Cancer.” The Analyst, vol. 135, no. 10, 2010, p. 2496, 10.1039/c0an00204f.

27. Fu, Ying, et al. Highly Sensitive Detection of Protein Biomarkers with Organic Electrochemical Transistors. Vol. 29, no. 41, 1 Nov. 2017, pp. 1703787–1703787, 10.1002/adma.201703787.

28. Li, Zhaohui, et al. “Rapid and Sensitive Detection of Protein Biomarker Using a Portable Fluorescence Biosensor Based on Quantum Dots and a Lateral Flow Test Strip.” Analytical Chemistry, vol. 82, no. 16, 19 July 2010, pp. 7008–7014, 10.1021/ac101405a.

29. Patil, Amol V, et al. “Graphene-Based Protein Biomarker Detection.” Bioanalysis, vol. 7, no. 6, 1 Apr. 2015, pp. 725–742, 10.4155/bio.15.4. Accessed 1 Aug. 2025.

30. Flores, Raja, et al. “Association of Stage Shift and Population Mortality among Patients with Non–Small Cell Lung Cancer.” JAMA Network Open, vol. 4, no. 12, 17 Dec. 2021, p. e2137508, http://www.ncbi.nlm.nih.gov/pmc/articles/PMC8683966/, 10.1001/jamanetworkopen.2021.37508.

31. Borrebaeck, Carl A. K. “Precision Diagnostics: Moving towards Protein Biomarker Signatures of Clinical Utility in Cancer.” Nature Reviews Cancer, vol. 17, no. 3, 1 Mar. 2017, pp. 199–204, http://www.nature.com/articles/nrc.2016.153, 10.1038/nrc.2016.153.

32. Schaaij-Visser, Tieneke B.M., et al. “Protein Biomarker Discovery for Head and Neck Cancer.” Journal of Proteomics, vol. 73, no. 10, Sept. 2010, pp. 1790–1803, 10.1016/j.jprot.2010.01.013.

33. Xiong, Hongjie, et al. “Cancer Protein Biomarker Discovery Based on Nucleic Acid Aptamers.” International Journal of Biological Macromolecules, vol. 132, 1 July 2019, pp. 190–202, http://www.sciencedirect.com/science/article/abs/pii/S0141813018373434, 10.1016/j.ijbiomac.2019.03.165.

34. American Cancer Society. “Survival Rates for Pancreatic Cancer.” http://www.cancer.org, 5 Feb. 2024, http://www.cancer.org/cancer/types/pancreatic-cancer/detection-diagnosis-staging/survival-rates.html.

35. Canva. “Canva.” Canva, 2025, http://www.canva.com/.

